# EHR-QC: A streamlined pipeline for automated electronic health records standardisation and preprocessing to predict clinical outcomes

**DOI:** 10.1101/2023.05.30.23290765

**Authors:** Yashpal Ramakrishnaiah, Nenad Macesic, Anton Y. Peleg, Sonika Tyagi

## Abstract

The adoption of electronic health records (EHRs) has created opportunities to analyze historical data for predicting clinical outcomes and improving patient care. However, non-standardized data representations and anomalies pose major challenges to the use of EHRs in digital health research. To address these challenges, we have developed EHR-QC, a tool comprising two modules: the data standardization module and the preprocessing module. The data standardization module migrates source EHR data to a standard format using advanced concept mapping techniques, surpassing expert curation in benchmarking analysis. The preprocessing module includes several functions designed specifically to handle healthcare data subtleties. We provide automated detection of data anomalies and solutions to handle those anomalies. We believe that the development and adoption of tools like EHR-QC is critical for advancing digital health. Our ultimate goal is to accelerate clinical research by enabling rapid experimentation with data-driven observational research to generate robust, generalisable biomedical knowledge.

**Highlights:**

- EHR-QC accepts EHR data from a relational database or as a flat file and provide an easy-to-use, customized, and comprehensive solution for data handling activities.
- It offers a modular standardization pipeline that can convert any EHR data to a standardized data model i.e. OMOP-CDM.
- It includes an innovative algorithmic solution for clinical concept mapping that surpasses the current expert curation process.
- We have demonstrated that the imputation performance depends on the nature and missing proportion, hence as part of EHR-QC we included a method that searches for the best imputation method for the given data.
- It also contains an end-to-end solution to handle other anomalies such as outliers, errors, and other inconsistencies in the EHR data.

## 1. Introduction

Electronic Health Records (EHR) have been widely adopted and contain an incredible wealth of digital health information including demographics, observations, investigations, diagnoses, treatments, procedures, and clinical notes. This has allowed EHR data to be used for many purposes, such as in public health surveillance [1, 2], disease modelling [3], predictive analytics [4, 5, 6], assessment of medical treatments and procedures [7], decision making [8] and policy development [9], and data-driven research [10, 11, 12]. These applications are possible because of the increased adoption of EHRs coupled with the emergence of data-driven machine-learning techniques that facilitate the ability to leverage large amounts of data to uncover hidden knowledge. However, there are significant limitations in use of EHR due to non-standardisation and inherent biases in the data [13, 14, 15, 16, 17].

One of the major challenges in conducting research using EHR data is the presence of anomalies, such as missing data, outliers, errors, and inconsistencies [18]. EHR data comprises various data types collected from different systems, some of which are obtained directly from monitoring devices, while others are entered manually [19]. Additionally, as the data is collected primarily for administrative purposes, it may not undergo the same level of rigorous vetting as manually collected research data leading to poor research outcomes when used unprocessed [20]. Even seemingly insignificant errors can have severe consequences, as evidenced by a study in which children whose weights were wrongly recorded resulted in drug overdose in one in three cases [21]. To handle these anomalies, several domain-specific frameworks [14, 20] and tools such as Achilles [22], DataQualityDashboard (DQD) [14], ARES, MIRACUM [23], mosaicQA [24], and Mind the gap [25] have been developed. However, many of these solutions are limited to a specific source format or scope, and do not offer a means to address identified anomalies. Therefore, it is necessary to create more effective quality control frameworks.

Furthermore, the lack of standardization of EHR data poses a significant challenge, especially with respect to data formatting, clinical terminology, analytical methods, and procedures. In our study, we prioritize two key aspects of standardization: data format and clinical terminology. Standardizing data format allows for seamless integration and analysis across different sources, while harmonizing clinical terminology facilitates accurate interpretation and comparison of findings [26, 27]. Conversely, non-standard data representation can negatively impact the adaptability of tools and methods to various sources leading to poor generalization of results, duplicated efforts, and laborious downstream processing [28, 29].

The EHR is typically stored in institution-specific databases, each with its own data representation format, also known as a “schema”. Standardizing the database schema involves maintaining a consistent data representation that are interoperable. To establish a standardized representation of EHR data, open-source common data models (CDMs) have been developed. The Observational Medical Outcomes Partnership-Common Data Models (OMOP-CDM), created by the OHDSI consortium, is one such example. Efforts are underway to convert custom schemas to the OMOP-CDM standard while preserving its content [30, 31, 32, 33, 34, 35, 36, 37, 38, 39]. This conversion allows for consistent data analysis and integration across different healthcare systems, facilitating collaborative research and improving interoperability [40] [41, 42].

However, the scope of these efforts is limited since they are tailored to a specific EHR format and cannot be repurposed. To address this issue, a few generic utilities have been developed such as the user-interface-based “White Rabbit”, and “Rabbit In A Hat” and purely code-based “Extract, transform, load (ETL) framework” [43] to facilitate the OMOP conversion. The user-interface-centric design of these tools makes them incompatible with version control systems, resulting in a need for manual intervention, scalability challenges, replication difficulties, and a higher risk of errors. In contrast, the code-based approach is less efficient, and its performance may not be sufficient for time-sensitive applications, particularly when dealing with large datasets.

The second challenge to EHR data standardisation is the use of free-text to document and store medical concept information such as clinical findings, procedures, and outcomes. To address this, clinical terminology standardization involves mapping clinical concepts to a standard vocabulary through a process known as concept mapping. In 2003, five controlled terminologies were proposed to represent different types of concepts, such as SNOMED-CT for clinical terms [44], LOINC for laboratory test orders and results [45, 46], RxNorm for clinical drugs [47], NDF RT for pharmacologic properties of medications, and UMDNS for medical devices [48](refer to supplementary table S1 for more details). These terminologies typically use hierarchical organizations and ontologies to define relationships between concepts. For example, an ontology may define the “is-a-type-of” relationship between “myocardial infarction” and “heart disease.” Repositories like UMLS [49] and Athena [50] provide access to these ontologies. However, the current process of mapping clinical concepts to standard ontologies is time-consuming and requires expert knowledge, even with tools like Usagi [32, 51, 52, 53]. Some concepts are particularly difficult to map, such as those in the drug exposure category, with only 38% being mapped in one study [32]. As a result, fully automating this process remains a significant challenge. The current best-performing methods involve an initial automated mapping followed by manual curation by experts. However, given the large number of medical concepts in an EHR dataset, manual mapping at scale is impractical in many cases. Overall, improving the quality and standardization of EHR systems is crucial to enhancing the scalability, reproducibility, and reliability of EHR-based research [54]. This would ultimately result in better healthcare quality, reduced costs, and wider adoption of EHR systems [55]. To tackle the above mentioned challenges, we have developed the EHR-QC toolkit. This fully-automated pipeline is specifically designed for the standardization of EHR data encoding, fully automated concept mapping, and comprehensive quality assurance of healthcare data. The EHR-QC toolkit has the potential to become an integral part of digital health workflows that rely on EHR data to perform observational studies.

## 2. Methods

The EHR-QC is composed of two main modules namely “Standardization Pipeline” (Figure 1.17) and the “Preprocessing Pipeline” (Figure 1.18) consisting of various utility Python code functions (Figure 1.1 - 1.16) for handling EHR. This toolkit is a command-line Python utility with a straightforward setup and user interface. A complete step-by-step guide to running the pipeline has been provided (https://ehr-qc-tutorials.readthedocs.io/). The following sections describe the technical details of different modules of the pipeline and provide case studies to demonstrate its utility.

### 2.1. Data sources

Two different data sources are used at different stages for developing and benchmarking EHR-QC modules. During the development of the standardization pipeline, we used the Medical Information Mart for Intensive Care (MIMIC) IV data, a de-identified EHR dataset collected from critical care settings in a US hospital [56]. Both the original MIMIC schema and its OMOP-CDM conversion were used to validate preprocessing functionalities. We also used a benchmark dataset obtained from a recent paper [57] on the conversion of UK Biobank EHR to OMOP-CDM. This dataset included concepts from three categories - “Operations,” “Non-Cancer Illnesses,” and “Cancers” - obtained from various EHR sources, along with their mappings to a standard ontology. The mappings were curated by a team of experts using Usagi tool in a semi-automated manner. The availability of expert-curated mappings was the primary reason for selecting this dataset as a benchmark, as it helps to ensure the accuracy and consistency of the concept mapping process.

### 2.2. Custom configuration setup

The pipeline is completely flexible to allow inputs both as an existing database schema and as flat text files in “.CSV” format containing any type and range of attributes. A collection of the module functions can be invoked as a single pipeline, such as the “Standardization Pipeline” (Figure 1.17) and the “Preprocessing Pipeline” (Figure 1.18) enabling complete automation of the end-to-end EHR data processing activity. Appropriate initial configurations are provided through a configuration file. The configuration file allows users to manage the customisations such as the database connection parameters, intermediate schema names for lookup, source, extract and, CDM tables, standard vocabulary file paths, paths of EHR source files, column mappings for each of these files, and boundary values for various attributes for performing data quality checks. These configuration options make the pipeline flexible by adapting to any variation in the source data and also to run in a fully automated manner. Detailed custom use cases are provided in our online documentation of the pipeline.

### 2.3. Migrating the EHR data to the OMOP-CDM schema

The OMOP-CDM migration module provides utility functions to facilitate the process of converting any EHR representation to the OMOP-CDM schema. The database templates for the migration scripts obtained from an earlier migration effort [58] are embedded within the Python codebase, which dynamically builds queries based on the configurations, forming a layer of abstraction for users. This module provides automatic end-to-end migration functionality of any EHR to the OMOP-CDM, including standard vocabulary import and concept mapping.

The first step in the migration process is to import the standard ontologies (Figure 1.5, Figure 1.6) and the HER (Figure 1.2) in their raw format into a database. The EHR data can be sourced either from a custom schema (Figure 1.1) or as a structured tabular flat file (Figure 1.4) typically formatted as CSV files. In this step, appropriate column mappings are to be provided if the data structure varies from the expected convention as shown in Figure S1. In the next step, the imported information is dumped into staging tables (Figure 1.2, Figure 1.6), from which the standard entities are extracted by the process known as ETL without affecting the raw data in the import tables. Depending on the source schema, extracting the OMOP-CDM entities might involve filtering information or merging attributes from staging tables. Mapping non-standard concepts in the source EHR to a standard ontology term known as concept mapping (Figure 1.8) is the most crucial and time-consuming step of the process. This step involves standardising non-standardised concepts in the EHR by either automatically mapping them with the desired standard ontologies or importing a pre-built custom mapping file. To facilitate concept mapping, we have developed a novel method to automatically perform this process, the details of which are discussed in the next section 2.4. Next, during the extract step (Figure 1.9), the EHR data is cleaned if needed, mapped to concepts available in the vocabulary tables, and OMOP-CDM entities are extracted. In the final step, the extracted entities are unloaded (Figure 1.10) to the final OMOP-CDM database (Figure 1.11). In this module, all the intermediate tables are automatically created and stored for any further analysis and audit. Further, this enables individual stages to be run independently, also resuming from where it is left off when run in pipeline mode.

### 2.4. Mapping clinical concepts to controlled vocabulary

Concept mapping typically involves three scenarios. The first scenario is when the source data already adheres to the desired standard. In this case, data can be directly moved to the target schema after performing basic code integrity checks. The second scenario is when the source data is standardized using a different standard than the target ontology. For well-established and compatible standards, a pre-existing mapping can be used to obtain the corresponding desired standard ontology terms. However, when no readily available mapping exists, *de novo* mapping between the source EHR standard and the desired ontology standard must be performed. The third scenario is when the source contains concepts that are not mapped to any ontology and might be collected as free-flow text. In this case, concepts from the source EHR need to be mapped individually onto a standard ontology term. This is summarised in Supplementary Table S2

There are several techniques available for performing concept mapping by measuring the similarity of a search term to standard ontology terms. One such technique is approximate string matching, also known as fuzzy matching. Fuzzy matching provides a similarity score based on the Levenshtein distance [59] between two strings, which quantifies the character-level differences between them. This method works best when the source text is taken from a controlled vocabulary that has only minor variations from standard concepts. To increase flexibility when dealing with diverse vocabularies, fuzzy algorithms can be extended through reverse indexing, word tokenization, and conditional mapping of tokens. A very popular tool, Usagi (https://github.com/OHDSI/Usagi) developed by OHDSI consortium, uses an extended reverse indexing based technique internally to provide the matching terms. Next, semantic matching algorithms seek to find the closest meaning match between two phrases, instead of just comparing their textual composition. To accomplish this, word embeddings are generated, which are multi-dimensional distributed vectors representing each phrase. In an n-dimensional space, the embeddings of similar phrases are closer to each other while the embeddings of opposing phrases are more distant. Therefore, a possible match can be identified by selecting the standard concept whose embedding is closest to the embedding of the search phrase. The medical concept annotation tool Medcat used this technique to detect clinical concepts in texts [60]. Semantic matching takes into consideration the semantics of concepts, allowing for the mapping of a more generic and diverse terminology. However, it falls short of human-level performance, making standalone automatic mapping algorithms highly error-prone and not a viable alternative for semi-automatic expert curation. Basically, no single algorithm is the most effective in all scenarios, as their effectiveness depends on the nature of the data to be mapped, as depicted in Figure S2.

Standalone concept mapping techniques are not as effective as expert curation. This makes them unsuitable for complete automation. Therefore, we implemented Majority Voting, a composite approach that only retains mappings supported by more than one standalone algorithm. Although this approach improves mapping accuracy, it also reduces mapping coverage. To address this issue, we developed another composite algorithm called “Majority Voting Plus.” In the first step, this algorithm identifies all mappings supported by two or more algorithms, like Majority Voting. For the unmapped concepts, we use Medcat, Usagi, and Fuzzy in that order of preference to obtain the first available mapping. With this approach, we resolved the low coverage issue while retaining superior performance. We have included Majority Voting Plus as part of the standardisation module in EHR-QC, which provides the only fully automated solution for EHR standardisation to the best of our knowledge. The mappings can also be saved as a CSV file for manual review later.

### 2.5. Data preprocessing to perform exploratory analysis and the quality assurance

#### 2.5.1. Exploratory data analysis and reporting anomalies

The data preprocessing module is equipped with various functions that perform monotonous data preprocessing activities like extraction, exploration, quality assurance (QA), correction, and preparation of EHR data for downstream analysis tasks. The *extract* function can be invoked to generate flat files by specifying connection details to the source repository stored as a relational database such as in SQL or postgres. Subsequent functions can be executed independently, decoupled from the data source, since they accept flat files as inputs. This module’s objective is to standardise the EHR data preprocessing process by providing a convenient library.

The extract module (Figure 1.12) fetches the demographic, vitals, and lab measurement information from the OMOP-CDM schema by default, additionally, it can also be configured to read the data in the MIMIC IV format and saves it in a csv file. Next, the *exploration* module (Figure 1.13) is used to generate reports aimed at providing a comprehensive overview of the healthcare data. The reports contain information about the attributes’ type, count, range, distribution, and summary statistics, along with information on anomalies such as missing values.

Further, the *QA* module (Figure 1.14) can be used to generate visualisations and statistics highlighting common anomalies such as missing data, outliers, errors, and other systematic inconsistencies. Additionally, this module not only identifies anomalies but also quantifies each category and offers remediation recommendations. For instance, the report displays the count and percentage of missing data and outliers. It also detects the presence of multiple data standards or distributions which can indicate data contamination, by obtaining the data modality. Lastly, to check the plausibility of systematic inconsistencies the distributions of the attributes are plotted against the predetermined boundary conditions and are visualised.

#### 2.5.2. Handing anomalies: missingness

The correction module (Figure 1.15 - *ehrqc*.*qc*.*Impute*) handles the tasks of dealing with the missing data. It performs a comparative analysis of various imputation algorithms such as mean imputation, median imputation, K-nearest neighbours (KNN) imputation, MissForest [61], Expectation Maximisation [62], and Multiple Imputation [63] are performed on the dataset. The imputation algorithms are applied to a random set of missing values with a specific percentage of missingness simulated artificially, and the root mean square error (RMSE) values are calculated to determine the best imputation strategy for the given dataset. This is repeated for all the algorithms and the RMSE in each case is compared to determine which algorithm works the best for the given data and the given proportion of missingness. The best-performing algorithm is applied automatically to impute missing values with the least RMSE score.

#### 2.5.3. Handling anomalies: outliers

The correction module (Figure 1.15 - *ehrqc*.*qc*.*Anomalies*) utilizes Item Response Theory (IRT) [64], an ensemble of unsupervised outlier detection algorithms to detect the outliers. This algorithm returns an ensemble score for every data point which is a combination of the outlier scores obtained from multiple unsupervised algorithms. This technique avoids the use of hard-set boundaries found in traditional outlier detection methods, which can lead to biased analysis and the removal of genuine data. This technique combines results from multiple unsupervised outlier detection algorithms to assign an overall anomaly score to each data point. Data points exceeding a threshold score are considered extreme values and excluded from further analysis. Lastly, this module (Figure 1.16) includes two functions for standardising and re-scaling the data which is essential for many machine learning tasks. These utility functions enable the user to generate a quality-assured data matrix that can be used to perform predictive modelling.

## 3. Results and discussion

### Evaluation of MIMIC-IV EHR data migration to OMOP-CDM reveals improved data quality and utility

To validate the migration process, we utilized our standardization pipeline to transfer MIMIC-IV EHR data to the OMOP-CDM schema. The process resulted in significant improvements in quality and utility of the EHR. For instance, the pipeline efficiently excluded any unusable patient entries that lacked subsequent entries. Moreover, a new data table was created during the migration process to store death information extracted from the admissions table, to increase the accessibility. The flow of data from the source schema to the OMOP-CDM through intermediate tables is illustrated in Figure 2. As part of the migration workflow, we have developed an automated concept mapping technique which will be explicated in the following section. This provides an opportunity to resolve discrepancies that may arise from the use of multiple units of measurement and harmonise redundant concepts. In the entire migration process, intermediate tables function as audit tables that ensure complete transparency (S3). It is possible to monitor the data that has been successfully migrated to the destination tables, as well as the data that has not been migrated due to various reasons such as inadequate quality or mapping failure. We successfully migrated 337,942 individuals, 2,435,481 visit occurrences, 468,919,408 measurements, and 9,331 death records from MIMIC-IV EHR to the OMOP-CDM structure in the process.

**Figure 1:**
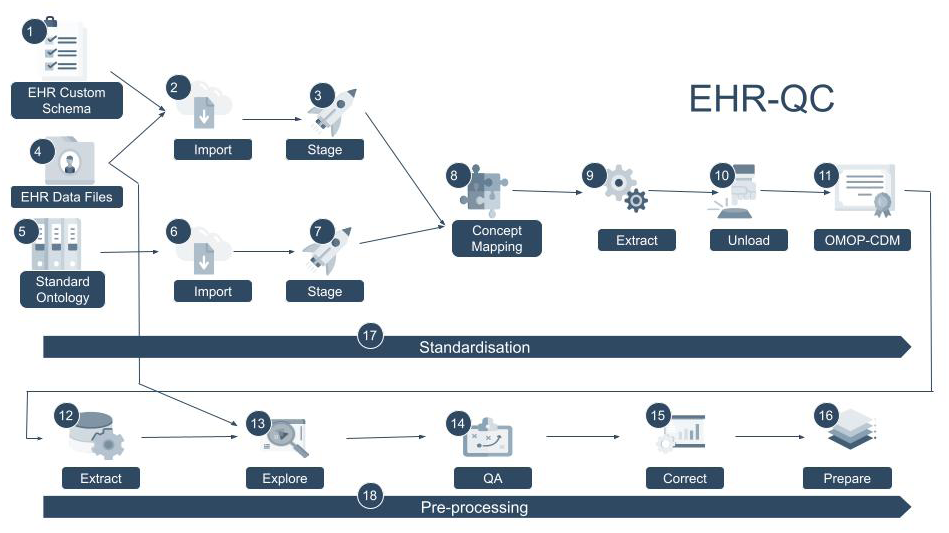
EHR-QC architecture diagram showing the standardisation and the preprocessing modules. The EHR data can be converted to OMOP-CDM standard using the standardisation module. This module imports and stages the data in its native format along with the reference vocabularies in an intermediate schema upon which the data is transformed into the standard entities. An important step in the process is to map the non-standard terms in the source data to a concept from the standard ontologies for which the pipeline provides an automated solution. This pipeline unloads the standardised data to the OMOP CDM schema as the final step. The EHR-QC also contains the preprocessing pipeline to assist with exploratory data analysis and quality assurance (QA) of the EHR data.

**Figure 2:**
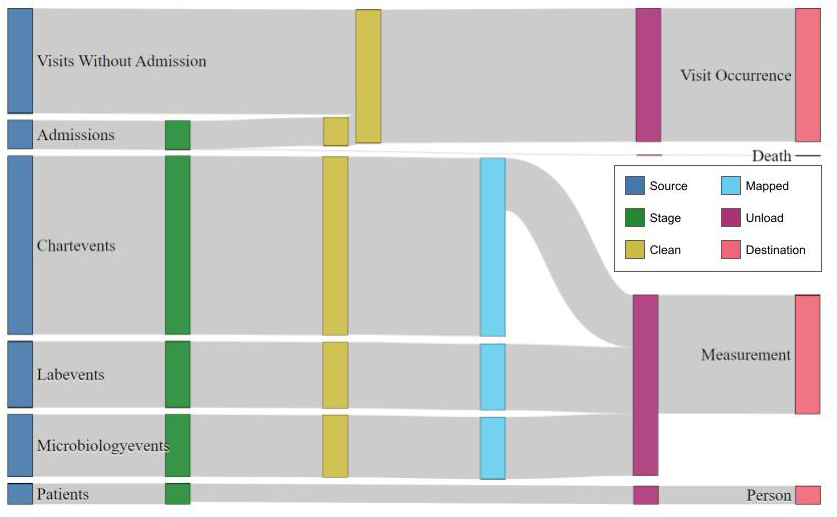
The figure illustrates the data flow diagram of four different entities. In all cases, the data from the source schema is imported into a staging table which is transformed into standard entities after performing cleaning and mapping to a desired standard. The standard entities are dumped in the unloading table which will be finally pushed to the destination OMOP-CDM schema.

### Majority Voting Plus outperforms expert-curated mappings with comprehensive coverage and high alignment

We benchmarked concept mapping performance of our pipeline using a published dataset [57]. Our results indicate that the Majority Voting approach yields a better alignment with curated concepts and minimises non-overlapping mappings when compared to standalone techniques (see Figure S3). However, since the Majority Voting gives a mapping only if there is a consensus amongst two or more standalone algorithms, it has a poor overall coverage as a substantial portion of the concepts remained unmapped due to the lack of consensus (see Figure 3A). The Majority Voting Plus technique on the other hand boosts coverage to a level similar to that of standalone algorithms (Figure 3A).

**Figure 3:**
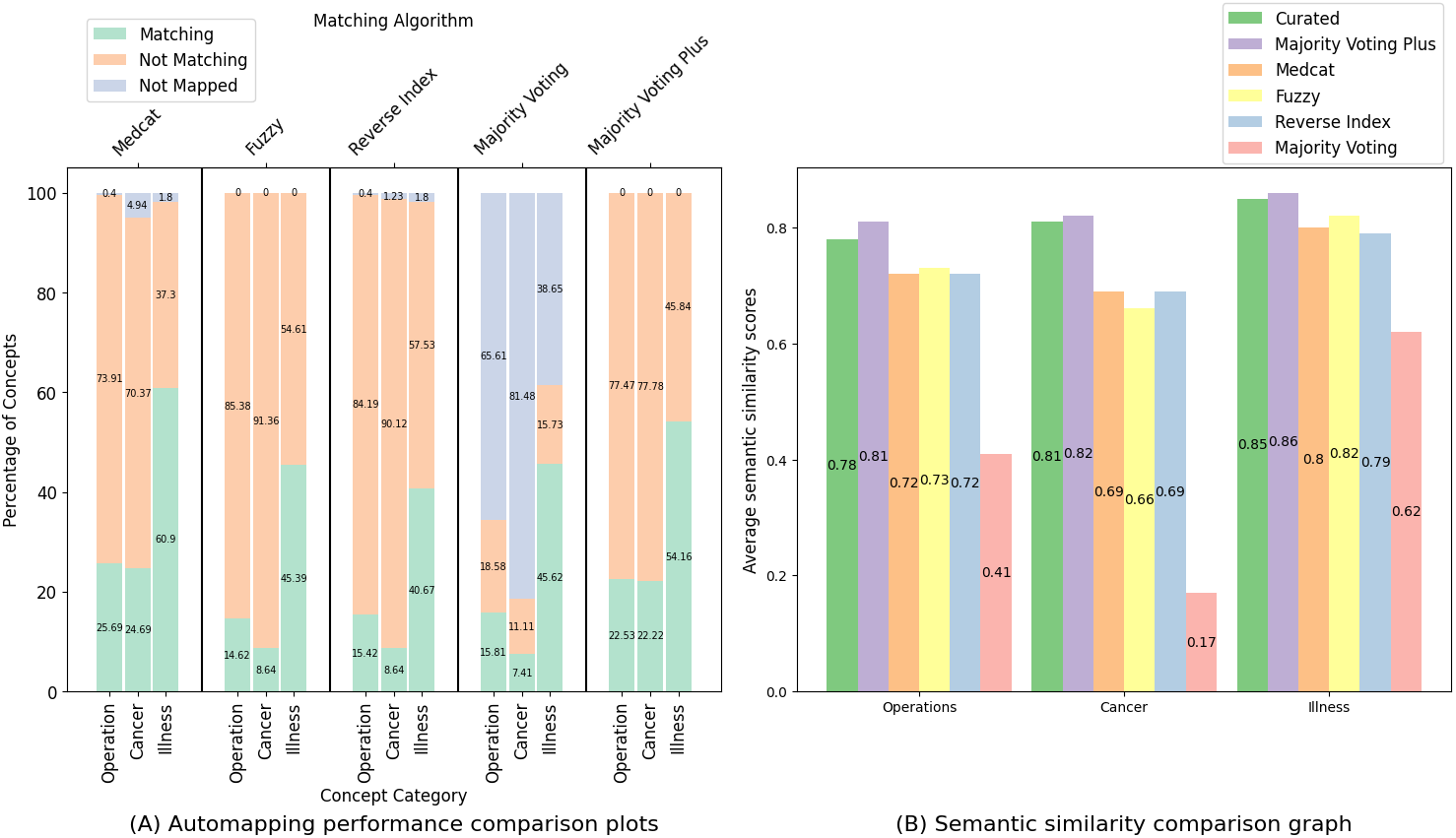
(A) An illustration of the percentage of coverage and mapping performance for Fuzzy, Reverse Index, Medcat, Majority Voting, and Majority Voting Plus techniques under three concept categories namely, Operation, Cancer, and Illness. The source texts that are not mapped are included under the Not Mapped category whereas the mappings that are in agreement with the expert-curated concepts are counted as Matching while the rest are included under Not Matching. (B) The average semantic similarity score which in this case is a population means taken for the measure of closeness in meaning to the source text. It is plotted for the expert-curated concepts along with the mappings from Fuzzy, Reverse Index, Medcat, Majority Voting, and Majority Voting Plus techniques. The medical concepts includes are Operations, Cancer, and Illness concept categories.

Further, we have evaluated the performance of our algorithm by comparing the mappings with the expert-curated concepts. The alignment between the two is presented in Figure (Figure 3A) where the proportions of mappings that match, do not match, or are not mapped to the curated values are displayed. The matching percentage gives the proportion of concepts in agreement with the curated concepts, providing a measure of the quality of the mapping. It is important to note that a non-matching percentage does not necessarily imply incorrectness, as both the curated and the mapped concepts may be correct in some cases, as shown in Supplementary Table S4. Our analysis demonstrates that concepts within the Illness category that are well-standardized exhibit a better alignment with curated mappings than those in other categories, as evidenced by the higher matching percentage of the algorithms. Conversely, the cancer concept type, which lacks a well-established standard vocabulary, performs poorly, particularly with text content matching algorithms like Fuzzy and Reverse Index. However, Medcat, a semantic matching technique, performs well in this category. Further optimization of Medcat’s performance can be achieved by fine-tuning it on more cancer-related text, as approximately 5% of the values remain unmapped. Our analysis also indicates that the Majority Voting Plus algorithm provides a mapping for every queried concept while maintaining a mapping percentage only slightly lower than that of Medcat. In summary, this figure shows that Majority Voting Plus and Medcat are the best-performing algorithms in terms of coverage and fidelity of the mappings.

Continuing our assessment of concept mapping, we utilised Semantic Similarity Score as a final criterion to measure the proximity of the mappings to the intended meaning of the concepts. Figure 3B compares the Semantic Similarity Scores between the query concepts and the mappings generated by the algorithms for three concept categories. The graphs show that expert-curated mappings were more similar to the query concepts than mappings derived from standalone techniques. However, our study found that the Majority Voting Plus approach consistently outperformed expert-curated mappings for all concept categories. This approach has comprehensive coverage and strong alignment with curated concepts, making it suitable for automatic concept mapping without compromising quality. This allowed us to fully automate the standardisation pipeline as part of the EHR-QC utility.

### Data exploration and quality reports provides an overview of the data and detect anomalies

We preprocessed EHR data in the standard OMOP-CDM format. Our first step was to generate data quality reports that contained exploration and anomaly graphs. Exploration graphs provided a comprehensive overview of the data, while anomaly graphs showed common anomalies such as missingness, outliers, inconsistencies, and systematic errors. The supplementary figure (Figure S4) presents the anomaly graphs generated by EHR-QC on the left, and the corresponding corrected data on the right. Our reports also included summary statistics of the data, such as the type and number of attributes, missingness and outliers, errors, and the proportion of data within user-specified value ranges. Overall, these reports are useful for gaining an overview of the EHR data and identifying anomalies.

### Missing data imputation simulates multiple imputation strategies and applies the best one for the given dataset

In addition to providing a general overview of the data and the anomalies, the EHR-QC preprocessing module includes utility functions to rectify the identified anomalies. Figure 4 presents the performance of missing data comparison and imputation utilities using various imputation techniques. In Plot 4A, the reconstruction r-squared score is plotted against different missing proportions ranging from 0 to 50 for various missing value imputation techniques. According to our analysis, the Expectation Maximisation algorithm performed the poorest among all the algorithms tested on the given data, regardless of the proportion of missing data. On the other hand, when the proportion of missing data ranged from 10-25%, MissForest showed the best performance, while for the 25-50% range, KNN outperformed the other algorithms. Interestingly, for missingness beyond 40%, Mean Imputation displayed the best performance. These findings demonstrate that the optimal algorithm for imputing missing data depends on the proportion of missing data in the dataset. Therefore, our results can guide the selection of the most appropriate algorithm based on the amount of missing data present in a given dataset.

**Figure 4:**
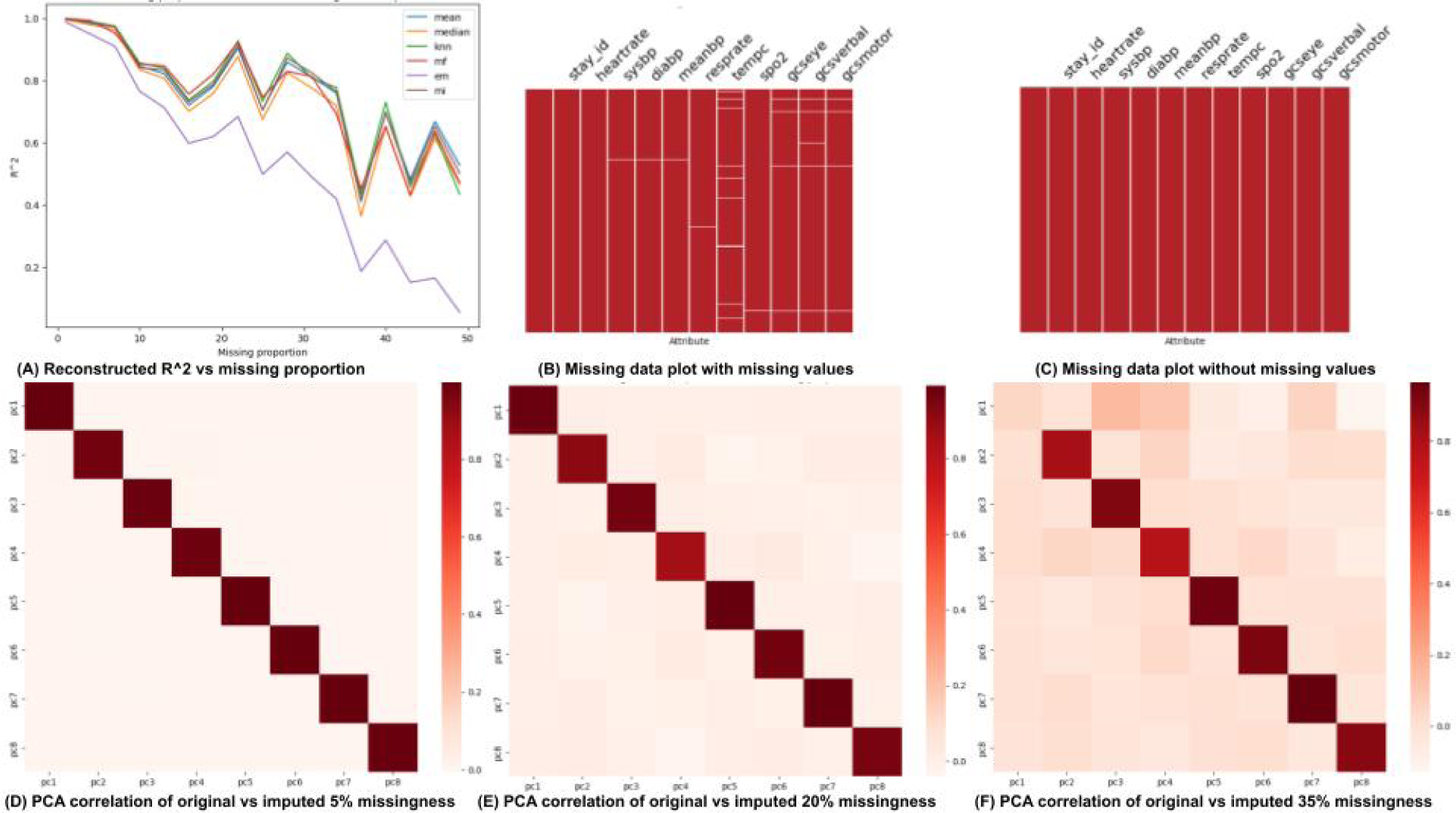
A) The reconstruction r squared values for some of the missing value imputation algorithms for a range of missing proportions. B) Missing data plots with missing values in the data. C) Missing data plots without missing values in the data. D) Correlation between the principal components of the original data and the imputed data from 5% missingness. E) Correlation between the principal components of the original data and the imputed data from 20% missingness. F) Correlation between the principal components of the original data and the imputed data from 35% missingness.

The plots 4B and 4C display the data with and without any missing values, respectively. Whereas, plots 4D, 4E, and 4F display the correlation between the principal components of the original data and the imputed data for simulated missingness of 5%, 20%, and 35%, respectively. These plots illustrate the performance degradation of the missing data imputation with the increase in missing data proportion highlighting the need for choosing an appropriate imputation technique suitable for the data provided.

### Outlier detection utility helps in the detection and treatment of outliers in an adaptive manner using an ensemble of unsupervised algorithms

We demonstrated how EHR-QC facilitates the identification and removal of outliers. Plots 5A, 5B and 5C in Figure 5 demonstrate EHR-QC’s ability to adaptively detect outliers. To demonstrate this, a simplified dataset with two attributes, “systolic blood pressure” and “heart rate,” was plotted in three scenarios. Plot 5A shows the unprocessed data, where extreme values are observed for both attributes. The ensemble score reaches as high as 25, indicating the presence of eccentric data points, represented by darker-colored points on the graph, which are extreme outliers. Plot 5B, obtained from the same dataset after applying a conventional univariate rule-based method to remove outliers, demonstrates the limitations of using inflexible hard cutoff values for classifying outliers. The boundaries imposed by this method result in the truncation of natural data clusters. Although this technique removed extreme outliers, a few data points with ensemble scores up to 10 remained. Plot 5C, obtained by applying an unsupervised method called IRT to identify outliers, demonstrates that the algorithm effectively removed outliers while retaining the entire cluster with its natural boundaries. The highest ensemble score for any data point in this plot did not exceed the value 6. Plots 5D, 5E, and 5F, demonstrating how EHR-QC plots aid in addressing inconsistencies in the data. Plot 5D shows the density plot of a single attribute, temperature, obtained from the raw data, indicating the presence of extreme values. After removing the outliers from this attribute, other anomalies now become apparent as shown in Plot 5E which in this case is the existence of multiple units of measurement. This plot uses vertical lines indicating normal ranges to aid in identifying such inconsistencies. Ideally, the majority of values should fall within the normal range, as shown in plot 5F, which was obtained after unifying the measurement standard.

**Figure 5:**
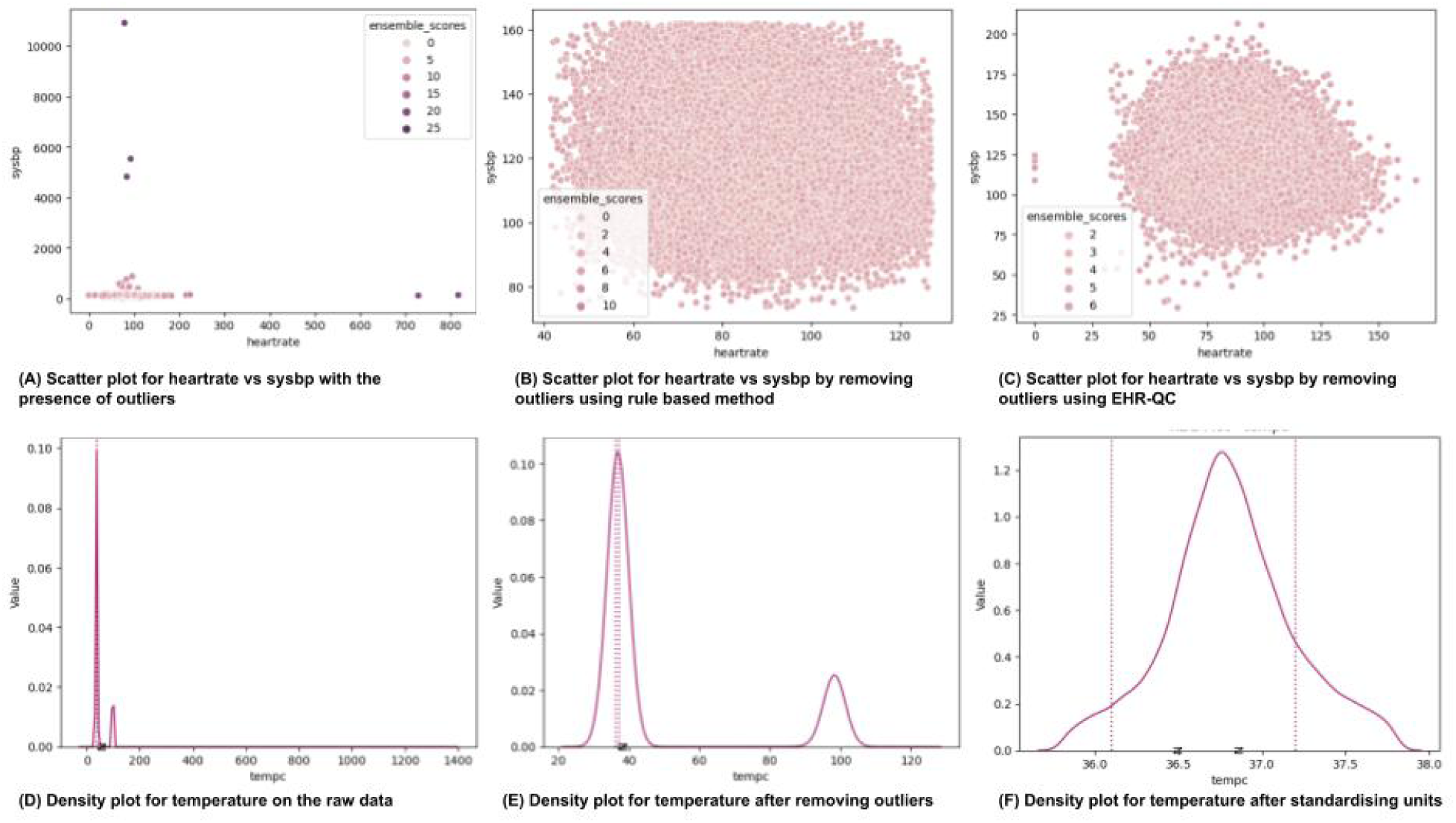
A) Scatter plot displaying ensemble outlier scores for systolic blood pressure vs heart rate plot for the raw dataset. B) Scatter plot displaying ensemble outlier scores for systolic blood pressure vs heart rate plot by removing outliers using a rule-based method. C) Scatter plot displaying ensemble outlier scores for systolic blood pressure vs heart rate plot by removing outliers using IRT. D) Density plot for the temperature attribute generated using the raw data. E) Density plot for the temperature attribute generated after removing the outliers. F) Density plot for the temperature attribute generated after removing the outliers and unifying the measurement standards.

## 4. Conclusion

Machine learning in digital health relies on large-scale healthcare data but is often limited to single-site data, hindering generalizability. Our work addresses this by providing a feasible EHR data harmonization workflow, enabling reproducible research outcomes through model validations on multi-site data.

Here, we introduced EHR-QC, a modular quality control pipeline that enables the conversion of EHR data to the standardized OMOP-CDM format. We also presented an innovative algorithmic solution for clinical concept mapping that surpasses the current expert curation process. This automation of data standardization represents a significant advancement, promoting the adoption of EHR standards and facilitating the development of more generalisable data-driven models. As a result, researchers can expect more efficient, reproducible, and robust research outcomes.

The EHR-QC pipeline also includes preprocessing functionalities for efficient exploration, quality assurance, and data preparation for downstream machine learning applications. Overall, EHR-QC offers a comprehensive and user-friendly solution for handling healthcare data, ensuring reproducible and robust research outcomes.

## Data Availability

The EHR-QC source code is now accessible to researchers for investigative purposes through the following Git repository: https://gitlab.com/superbugai/ehrqc.

## 5. Acknowledgements

We acknowledge contributions from two research interns, Esha Singh and Geeta Kole for their initial analysis of MIMIC to OMOP conversion workflow. We also thank Tyrone Chen for proofreading the manuscript, and Jerico Revote from Monash eResearch Centre for his invaluable support.

AP, NM, ST acknowledge Medical Research Future Fund funding for the SuperbugAI flagship project. YR received Monash Graduate Scholarship for his PhD.

The authors also extend their sincere appreciation to the open research community responsible for making the following resources available which were instrumental in facilitating the execution of this research; IRT [64], MIMIC IV [56], MIMIC IV to OMOP CDM Conversion (https://github.com/OHDSI/MIMIC), UK Biobank transformation to OMOP-CDM [57], Athena (https://athena.ohdsi.org/), SNOMED (https://www.snomed.org/), Usagi (https://github.com/OHDSI/Usagi), Medcat [60], and HuggingFace [65].

## 6. Code availability

The EHR-QC source code is now accessible to researchers for investigative purposes through the following Git repository: https://gitlab.com/superbugai/ehrqc. Comprehensive documentation for the utility can also be found at https://ehr-qc-tutorials.readthedocs.io.

## CRediT authorship contribution statement

**Yashpal Ramakrishnaiah:** Formal analysis, Investigation, Visualisation, Writing – review and editing, Software Development. **Nenad Macesic:** Investigation, Funding Acquisition, Supervision, Writing – review and editing. **Anton Y. Peleg:** Investigation, Funding Acquisition, Resources, Supervision, Writing – review and editing. **Sonika Tyagi:** Conceptualisation, Formal analysis, Investigation, Funding Acquisition, Resources, Supervision, Writing – review and editing.

