## Supplementary File for "EHR-QC: A streamlined pipeline for automated electronic health records standardisation and preprocessing to predict clinical outcomes"

| Acronym | Ontology | Description |
|---|---|---|
| SNOMED-CT | Systematized Nomenclature of Medicine - Clinical Terms | A systematically interpretable polyhierarchical subtype multi-lexical clinical terminology system |
| LOINC | Logical Observation Identifier Names and Codes | A Universal Standard for Identifying Laboratory Observations |
| RxNorm | Medical prescription normalised | A standardised nomenclature for clinical drugs |
| NDF RT | National Drug File - Reference Terminology | A concept-oriented terminology, a collection of concepts, each of which represents a single, unique meaning |
| UMDNS | Universal Medical Device Nomenclature System | A standard international nomenclature and computer coding system for medical devices |
| ICD | International Classification of Diseases | A globally used diagnostic tool for epidemiology, health management and clinical purposes |
| MeSH | Medical Subject Headings | A controlled and hierarchically-organised vocabulary used for indexing, cataloguing, and searching of biomedical and health-related information |
| CPT4 | Current Procedural Terminology | A numeric coding system consisting of descriptive terms and identifying codes |
| dm+d | Dictionary of medicines and devices | A dictionary of descriptions and codes which represent medicines and devices |
| EXACT | Experimental actions | A generic semantic representation of experimental protocols to ensure their reproducibility |

**Table S1**
Details of prominent standard ontologies.

| Scenario No | Source Standard | Required Standard | Mapping Available | Action |
|---|---|---|---|---|
| Scenario 1 | ICD 9 | ICD 9 | N/A | Use the source concept codes |
| Scenario 2A | ICD 9 | ICD 10 | Yes | Use the available mapping to get desired standard from source concepts |
| Scenario 2B | ICD 9 | NDC | No | Obtain *de novo* mappings and then use it to get desired standard from source concepts |
| Scenario 3 | None | NDC | N/A | Obtain desired standard concepts for the source concepts from the EHR individually |

**Table S2**
Table with different scenarios encountered during the mapping and their action points.

```
procedures = {
    'file_name': '/path/to/procedures_icd.csv',
    'column_mapping': {
        'subject_id': 'Subject ID column in procedures_icd.csv',
        'hadm_id': 'Hospital Admission ID column in procedures_icd.csv',
        'seq_num': 'Sequence Number column in procedures_icd.csv',
        'chartdate': 'Chart Date column in procedures_icd.csv',
        'icd_code': 'ICD code column in procedures_icd.csv',
        'icd_version': 'ICD version column in procedures_icd.csv',
    },
}
```

**Figure S1:** A sample configuration showing the file path and the column mappings for procedure entity

| | | Source | | Intermediate Tables | | Destination |
|---|---|---|---|---|---|---|
| Item ID | Vitals | Chartevents Count | Chartevents Clean Count | Chartevents Mapped Count | CDM Measurement Count | Measurement Count |
| 220045 | heartrate | 6798187 | 6798187 | 6798187 | 6798187 | 7486283 |
| 220050 | sysbp | 2379566 | 2379566 | 2379566 | 2379566 | 2612690 |
| 220179 | sysbp | 4279569 | 4279569 | 4279569 | 4279569 | 4719582 |
| 220051 | diasbp | 2379199 | 2379199 | 2379199 | 2379199 | 2612247 |
| 220180 | diasbp | 4278628 | 4278628 | 4278628 | 4278628 | 4718512 |
| 220052 | meanbp | 2387853 | 2387853 | 2387853 | 2387853 | 2622569 |
| 220181 | meanbp | 4276928 | 4276928 | 4276928 | 4276928 | 4716923 |
| 225312 | meanbp | 242870 | 242870 | 242870 | 242870 | 259934 |
| 220210 | resprate | 6728530 | 6728530 | 6728530 | 6728530 | 7410589 |
| 224688 | resprate | 363651 | 363651 | 363651 | 363651 | 400769 |
| 224689 | resprate | 629555 | 629555 | 629555 | 629555 | 683287 |
| 224690 | resprate | 583780 | 583780 | 583780 | 583780 | 632923 |
| 223761 | tempc | 1595844 | 1595844 | 1595844 | 1595844 | 1756560 |
| 223762 | tempc | 277621 | 277621 | 277621 | 277621 | 297934 |
| 220277 | SpO2 | 6656949 | 6656949 | 6656949 | 6656949 | 7327432 |
| 220739 | gcseye | 1711688 | 1711688 | 1711688 | 1711688 | 1877899 |
| 223900 | gcsverbal | 1708459 | 1708459 | 1708459 | 1708459 | 1875141 |
| 223901 | gscmotor | 1704243 | 1704243 | 1704243 | 1704243 | 1870608 |

**Table S3**
Counts of vitals through the migration process. Some data were combined or aggregated into non-redundant standard tables. For instance, various event tables at the source such as *chartevents*, *labevents*, and *microbiologyevents* were merged into a single table in the destination called *measurements*.

| Source Code | Source Name | Curated Concept | Medcat Concept |
|---|---|---|---|
| Curated concepts and the Medcat mappings are equally correct | | | |
| 1613 | ct colonoscopy | Virtual CT colonoscopy | Computed tomography of colon |
| 1250 | bell's palsy/facial nerve palsy | Bell's palsy | Facial palsy |
| 1583 | ischaemic stroke | Ischemic stroke | Cerebral infarction |
| Medcat mappings are closer in meaning to the source concepts than to the curated concepts | | | |
| 1595 | pleural plaques (not known asbestosis) | Asbestos-induced pleural plaque | Pleural plaque |
| 1279 | eye trauma | Traumatic injury | Contusion of eye |
| 1288 | nervous breakdown | Broken skin | Dysthymia |
| Curated concepts are closer in meaning to the source concepts than to the Medcat mappings | | | |
| 1682 | benign insulinoma | Benign insulinoma | Benign |
| 1066 | heart/cardiac problem | Heart disease | Heart structure |
| 1408 | alcohol dependency | Alcohol dependence | Alcohol |

**Table S4**
Table containing selected examples of source concepts along with the curated and Medcat mappings for three different scenarios.

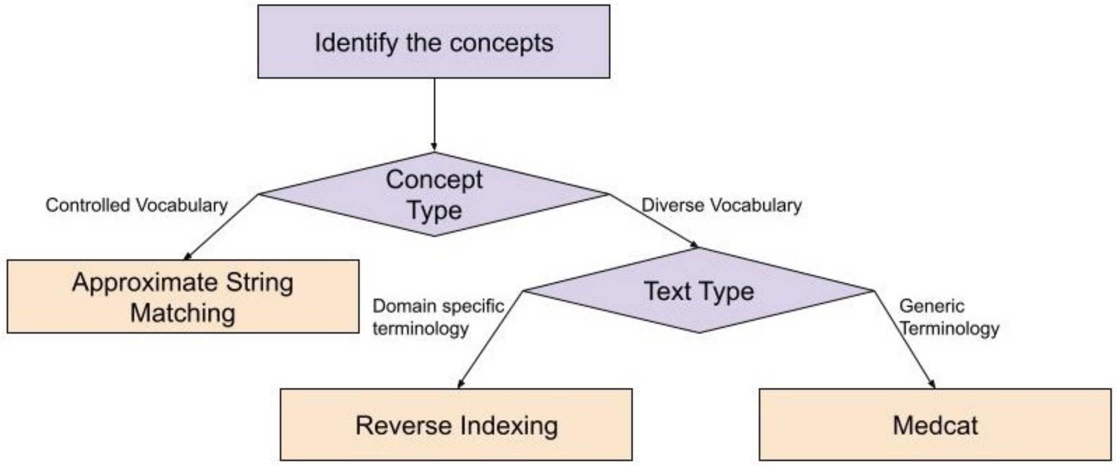


**Figure S2:** A flow chart for choosing the right concept mapping strategy. The first step in this process is to extract the concepts to be mapped from the source data. Next, if the terminology in the concept to be mapped is obtained from a controlled vocabulary like the drug names, a simple fuzzy text matching technique such as "Approximate String Matching" will be sufficient. On the other hand, if it resembles the free flow text, it is needed to determine if the terminology used is domain specific or generic in nature. Structured search tools like Usagi perform well if the terminology is consistent with the standard to be mapped such as clinical procedures or observations. Whereas, a much complex and computationally intensive method such as Medcat which is based on semantic mapping will be necessary if the concepts consist of generic terminology as in the admission and discharge locations.

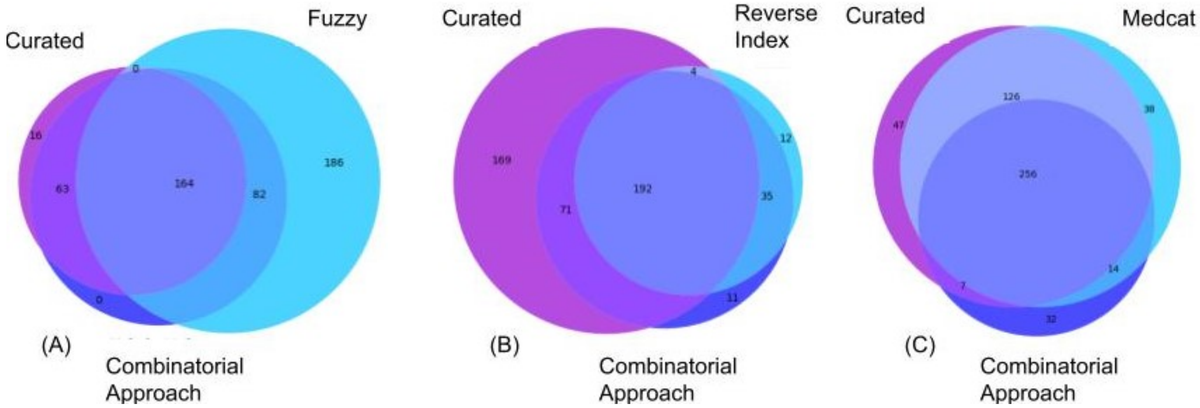


**Figure S3:** A) Diagram showing overlap between curated concepts, mapped concepts obtained by consensus-based combinatorial approach, and mapped concepts obtained by Fuzzy algorithm B) Diagram showing overlap between curated concepts, mapped concepts obtained by consensus-based combinatorial approach, and mapped concepts obtained by Reverse Index algorithm C) Diagram showing overlap between curated concepts, mapped concepts obtained by consensus-based combinatorial approach, and mapped concepts obtained by Semantic mapping algorithm.

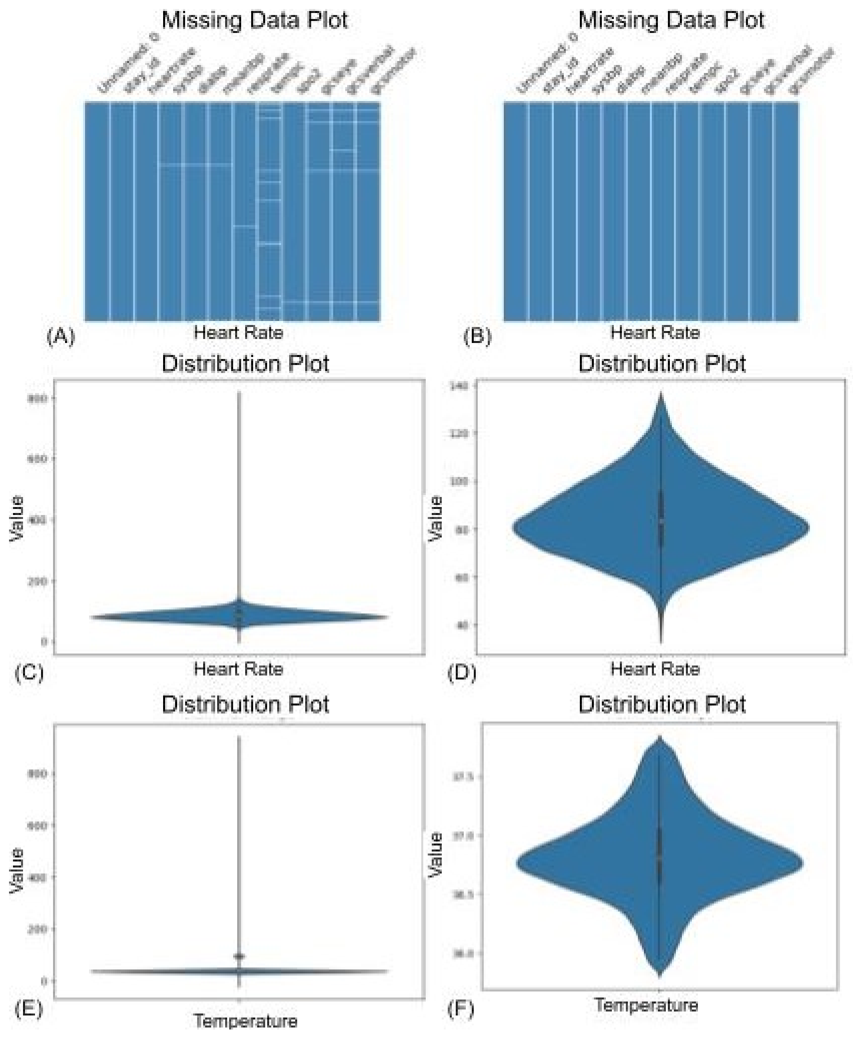


**Figure S4:** A) Missing data plots with missing values in the data. B) Missing data plots without missing values in the data. C) Violin plot showing the distribution of the heart rate before removing the outliers. D) Violin plot showing the distribution of the heart rate after removing the outliers. E) Violin plot showing the distribution of the temperature before standardisation and outlier removal. F) Violin plot showing the distribution of the temperature after standardisation and outlier removal.
